# Impact of Advanced Therapy Centers on Characteristics and Outcomes of Heart Failure Admissions

**DOI:** 10.1101/2023.08.07.23293806

**Authors:** Daniel Y Lu, Jaya Kanduri, Ilhwan Yeo, Parag Goyal, Udhay Krishnan, Evelyn M Horn, Maria G Karas, Irina Sobol, David T Majure, Yoshifumi Naka, Robert M Minutello, Jim W Cheung, Nir Uriel, Luke K Kim

## Abstract

**Background:** Although much attention has been paid to admission and transfer patterns for cardiogenic shock, contemporary data is lacking on decompensated heart failure (HF) admissions and transfers, and the impact of advanced therapy centers (ATCs) on outcomes.

**Methods:** HF hospitalizations were obtained from the Nationwide Readmissions Database 2016-2019. Centers performing at least one heart transplant or left ventricular assist device were classified as ATCs. Patient characteristics, outcomes, and procedural volume were compared among three cohorts: admissions to non-ATCs, admissions to ATCs, transfers to ATCs. A secondary analysis evaluated outcomes for severe HF hospitalizations (cardiogenic shock, cardiac arrest, mechanical ventilation).

**Results:** 2,331,690 hospitalizations were admissions to non-ATCs (94.5% of centers), 525,037 were admissions to ATCs (5.5% of centers), and 15,541 were transfers to ATCs. Patients treated at ATCs (especially those transferred) had higher rates of HF decompensations, procedural frequency, lengths-of-stay, and costs. Unadjusted mortality was 2.6% at non-ATCs and was higher at ATCs, both for directly admitted (2.9%, p<0.01) and transferred (11.2%, p<0.01) patients. However, multivariable adjusted mortality was significantly lower at ATCs, both for directly admitted (OR 0.82, p<0.01) and transferred (OR 0.66, p<0.01) patients. For severe HF admissions, unadjusted mortality was 37.2% at non-ATCs and was lower at ATCs, both for directly admitted (25.3%, p<0.01) and transferred (25.2%, p<0.01) patients, with similarly lower multivariable adjusted mortality.

**Conclusions:** HF patients treated at ATCs were sicker but associated with higher procedural volume and lower adjusted mortality.

**Clinical Perspective:** Contemporary data is lacking on admissions and transfers for decompensated heart failure (HF) and the impact of advanced therapy centers (ATCs) on outcomes. Our findings show that decompensated HF patients treated at ATCs had higher rates of HF decompensations, procedural frequency, lengths-of-stay, and costs. While unadjusted mortality was higher at ATCs, multivariable adjusted mortality was significantly lower at ATCs, both for directly admitted and transferred patients. Our findings will hopefully prompt earlier recognition and referral of patients to ATCs, emphasize the need for increased numbers of ATCs, and spark further research into the decision-making process for referral to ATCs.

## Introduction

The burden of heart failure (HF) has been rising in the United States, with up to 6.2 million adults currently diagnosed with HF^1^, 1 million new diagnoses per year^2^, and a projected 46% increase in prevalence by 2030^3^. Primary HF hospitalization rates have been increasing, from 4.2 per 1000 US adults in 2014 up to 4.9 in 2017^4^. With rising burden comes cost, with over $30 billion due to HF in 2012, and a projected rise to $70 billion by 2030^1,3^. Despite advancements of medical and device therapy, HF remains associated with significant mortality, with over 40% of patients dying within 5 years^3^. Hospitalizations for decompensated HF particularly portend poor outcomes^2,3,5^. Despite aggressive management, many patients have progression of disease warranting advanced therapies such as durable left ventricular assist devices (LVAD) or heart transplant^5^, which can only take place at specialized “advanced therapy centers” (ATCs).

In recent years, focus has been garnered to early recognition and intensive therapy for cardiogenic shock, the most severe manifestation of HF. Many hospital systems have successfully developed protocols, teams, and networks to coordinate early referral/transfer to hub hospitals and subsequent therapies, with improved outcomes^6–11^. On the other hand, despite its morbidity, many all-comer HF patients are only seen by internal medicine, without timely referral to appropriate specialists or capable centers^12^. Although ATCs represent only a small minority of hospitals that treat decompensated HF, and only a small proportion of patients ultimately require advanced therapies, ATCs nevertheless can offer additional resources, specialists, interventions, and therapies that have the potential to improve outcomes for HF of all severity. This study seeks to use a large, nationally representative database to evaluate the impact of ATCs on the characteristics and outcomes of HF admissions and transfers.

## Methods

### Data Source

Data for the study were obtained from the 2016-2019 Nationwide Readmissions Database (NRD). The NRD is maintained by the Agency for Healthcare Research and Quality (AHRQ) through the Healthcare Cost and Utilization Project (HCUP). The NRD is a nationally representative database drawn from the HCUP State Inpatient Databases (SID), utilizing individual calendar years of discharge data, with patient linkage numbers to track patients across hospitalizations in a given year within a state. In 2019, the NRD contained deidentified information for over 18 million discharges from over 2,500 hospitals across 30 states, representing over 35 million weighted discharges^13^. Patient records in the NRD contain information about the hospitalization through diagnoses and procedures codes based on the International Classification of Diseases, Tenth Revision–Clinical Modification (ICD-10-CM) codes. Given all data were derived from a de-identified national administrative database, Institutional Review Board (IRB) approval and informed consent were not required. The data that support the findings of this study are available from the corresponding author upon reasonable request, and all data and materials have been made publicly available at the AHRQ.

### Advanced Therapy Centers and Interhospital Transfer

LVAD implantation and heart transplant surgeries were identified utilizing ICD-10-CM procedure codes (Supplemental Table 1). Centers performing at least one LVAD or heart transplant in a given year were identified as ATCs; other centers were designated as non-ATCs. Interhospital transfers were identified using the variable samedayevent, while transfers to rehab centers were eliminated using the variable rehabtransfer. For such transfers, the NRD combines individual records from sending and receiving hospitals into a single combined labeled hospitalization record^13^. Data were included only from hospitalizations for which there was a clear transfer or no-transfer designation.

### Study Population and Variables

Hospitalizations for patients age ≥ 18 were selected based on the presence of ICD-10-CM codes for HF as the primary diagnosis code (Supplemental Table 1). The first HF admission for a patient in a given year was identified as the index admission. To allow readmission analysis, index admissions occurring in the last 90 days of the year were excluded. ICD-10-CM codes for specific HF decompensations are listed in Supplemental Table 1, such as cardiogenic shock, cardiac arrest, mechanical ventilation, noninvasive ventilation >24 hours, ventricular arrhythmias, and acute kidney injury. Hospitalizations were separated into three cohorts – (A) direct admission to non-ATC, (B) direct admission to ATC, (C) transfer to ATC. Hospitals and patients from each year were considered separate entities, since the NRD does not provide year-to-year linkage of data

Hospital- and patient-level characteristics, comorbidities, outcomes, and heart-failure related procedures were identified using NRD variables, AHRQ comorbidity measures, and ICD-10-CM codes (Supplemental Table 1). The primary outcome was in-hospital mortality. Secondary outcomes include HF-related procedural frequency, estimated cost, lengths-of-stay, and 90-day readmission in survivors. Patients that died during their index hospitalization were excluded from subsequent readmission analyses.

After initial analysis, a secondary analysis was performed similarly to further evaluate the characteristics and outcomes of hospitalizations for severe HF, where the first index HF admission involved the most severe/morbid HF decompensations, as defined by cardiogenic shock, cardiac arrest, or mechanical ventilation.

### Statistical Analysis

Analyses were performed using SAS, version 9.4 (SAS Institute, Cary, NC). NRD discharge weights were used to calculate national estimates^14^. For comparison of categorical variables, the Rao-Scott χ2 test was used, and for continuous variables, survey-specific linear regression was used, both accounting for complex survey design. Multivariable logistic regression analyses were used to analyze the association of index hospitalization and patient characteristics, including presence of HF decompensations, with admission to ATC or transfer to ATC. To estimate cost for each hospitalization, the charge for each hospitalization was multiplied by the respective cost-to-charge ratios provided separately by HCUP. Statistical tests used were 2-sided, with p < 0.01 representing statistical significance.

## Results

### Characteristics of Hospitals and Patients

The majority of hospitals treating HF were non-ATCs (94.5%), while a minority were ATCs (5.5%) (Central Illustration). All ATCs were metropolitan, with the vast majority being large, teaching hospitals (Supplemental Table 2). For all index HF hospitalizations, 2,331,690 (81.2%) were direct admissions to non-ATCs, 525,037 (18.3%) were direct admissions to ATCs (Cohort B), and 15,541 (0.5%) were transfers to ATCs (Central Illustration, Table 1). Notably, nearly 19% of all HF hospitalizations were treated at the 5.5% of ATCs.

**Figure 1–.**
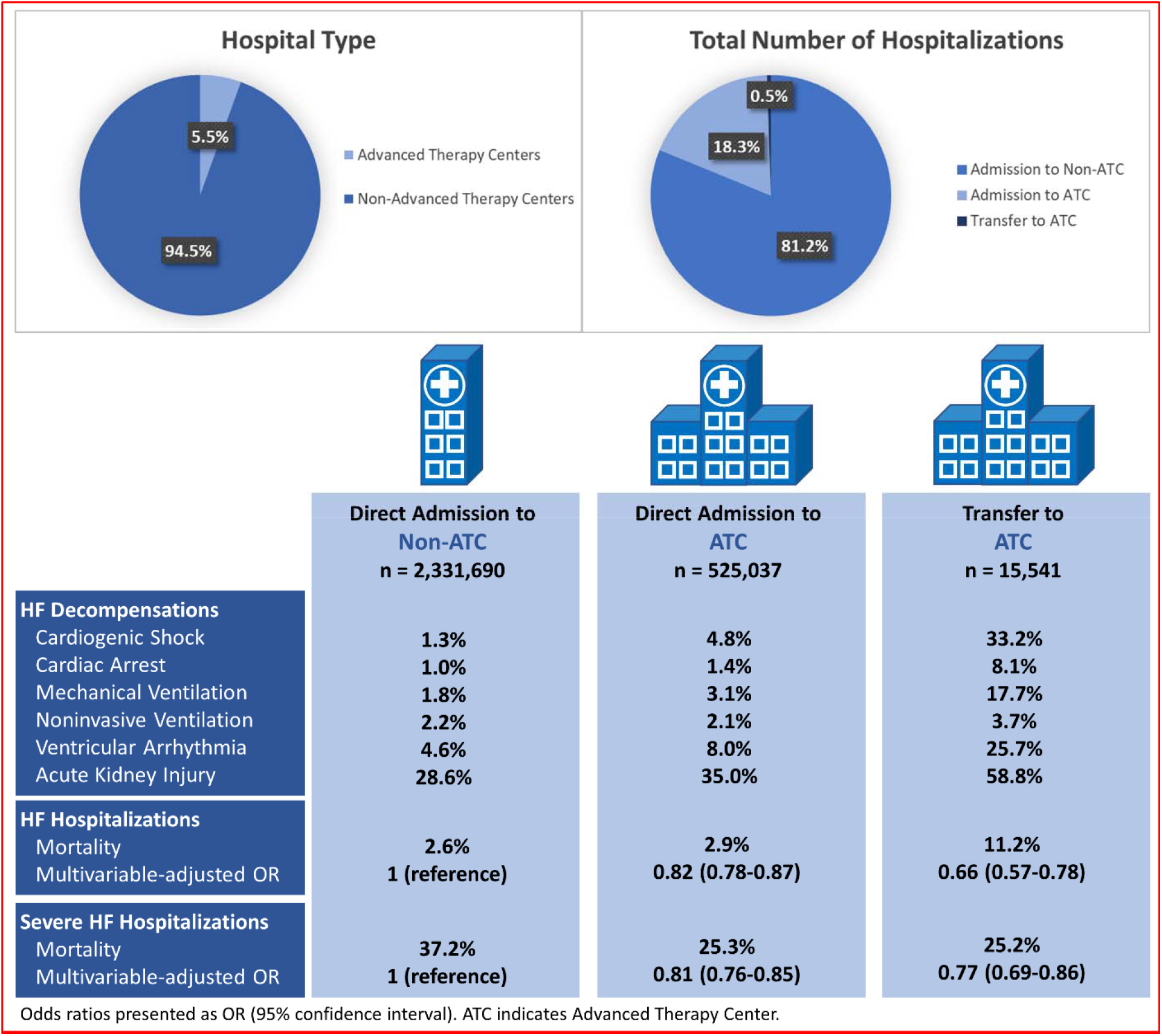
Hospital Types, Characteristics, and Outcomes for Heart Failure Hospitalizations 2016-2019

**Table 1-.** Characteristics of Patients Admitted with Heart Failure, Stratified by Hospital Type and Transfer Status.

| **Characteristic** | **Admission to**<br>**Non-ATC**<br>**(A)**<br>n = 2,331,690 | **Admission to**<br>**ATC**<br>**(B)**<br>n = 525,037 | **P value\*** | **Transfer to**<br>**ATC**<br>**(C)**<br>n = 15,541 | **P value\*** |
| --- | --- | --- | --- | --- | --- |
| **Demographics** |  |  |  |  |  |
| Age, mean | 72.5<br>(SEM = 0.06) | 69.3<br>(SEM = 0.19) | <0.01<sup>†</sup> | 63.2<br>(SEM = 0.29) | <0.01<sup>†</sup> |
| Age, group |  |  |  |  |  |
| <40 | 54,237 (2.3%) | 21,365 (4.1%) | <0.01 | 1,345 (8.7%) | <0.01 |
| 40-49 | 109,894 (4.7%) | 33,871 (6.5%) |  | 1,462 (9.4%) |  |
| 50-59 | 269,154 (11.5%) | 76,594 (14.6%) |  | 2,816 (18.1%) |  |
| 60-69 | 444,271 (19.1%) | 114,752 (21.9%) |  | 4,198 (27.0%) |  |
| 70-79 | 576,446 (24.7%) | 122,629 (23.4%) |  | 3,516 (22.6%) |  |
| 80+ | 877,688 (37.6%) | 155,826 (29.7%) |  | 2,204 (14.2%) |  |
| Female sex | 1,147,608 (49.2%) | 245,193 (46.7%) | <0.01 | 6,152 (39.6%) | <0.01 |
| Median household income<br>by zip code (quartile) |  |  |  |  |  |
| 1<sup>st</sup> | 758,151 (33.0%) | 172,694 (33.2%) | <0.01 | 3,955 (25.8%) | <0.01 |
| 2<sup>nd</sup> | 651,340 (28.3%) | 126,395 (24.3%) |  | 4,427 (28.9%) |  |
| 3<sup>rd</sup> | 527,244 (22.9%) | 124,017 (23.9%) |  | 3,905 (25.5%) |  |
| 4<sup>th</sup> | 363,826 (15.8%) | 96,611 (18.6%) |  | 3,040 (19.8%) |  |
| **Comorbidities** |  |  |  |  |  |
| Obesity | 605,217 (26.0%) | 137,546 (26.2%) | 0.51 | 4,693 (30.2%) | <0.01 |
| Hypertension | 666,827 (28.6%) | 148,903 (28.4%) | 0.86 | 7,952 (51.2%) | <0.01 |
| Hyperlipidemia | 1,247,109 (53.5%) | 276,375 (52.6%) | 0.19 | 8,879 (57.1%) | <0.01 |
| Coronary Artery Disease | 1,286,280 (55.2%) | 292,342 (55.7%) | 0.27 | 10,898 (70.1%) | <0.01 |
| Prior Myocardial Infarction | 333,104 (14.3%) | 77,044 (14.7%) | 0.26 | 2,839 (18.3%) | <0.01 |
| Atrial Fibrillation/Flutter | 1,050,607 (45.1%) | 231,321 (44.1%) | 0.02 | 7,415 (47.7%) | <0.01 |
| Valvular Disease | 670,882 (28.8%) | 159,524 (30.4%) | <0.01 | 7,650 (49.2%) | <0.01 |
| Peripheral vascular disease | 254,419 (10.9%) | 55,024 (10.5%) | 0.06 | 2,406 (15.5%) | <0.01 |
| Diabetes | 1,101,202 (47.2%) | 245,002 (46.7%) | 0.13 | 6,991 (45.0%) | <0.01 |
| Chronic Kidney Disease | 1,162,466 (49.9%) | 278,921 (53.1%) | <0.01 | 8,165 (52.5%) | <0.01 |
| Liver Disease | 132,891 (5.7%) | 40,931 (7.8%) | <0.01 | 2,743 (17.7%) | <0.01 |
| Malignancy | 117,022 (5.0%) | 29,543 (5.6%) | <0.01 | 934 (6.0%) | <0.01 |
| Thyroid Disorder | 457,759 (19.6%) | 96,236 (18.3%) | <0.01 | 3,148 (20.3%) | 0.26 |
| Chronic pulmonary disease | 909,008 (39.0%) | 175,158 (33.4%) | <0.01 | 5,741 (36.9%) | <0.01 |
| Autoimmune Conditions | 83,051 (3.6%) | 20,385 (3.9%) | <0.01 | 779 (5.0%) | <0.01 |
| Malnutrition/Weight loss | 130,743 (5.6%) | 38,028 (7.2%) | <0.01 | 2,394 (15.4%) | <0.01 |
| Coagulopathy | 167,706 (7.2%) | 47,983 (9.1%) | <0.01 | 3,413 (22.0%) | <0.01 |
| Long term anticoagulation | 520,313 (22.3%) | 119,395 (22.7%) | 0.31 | 3,982 (25.6%) | <0.01 |
| Tobacco Use | 982,779 (42.1%) | 211,688 (40.3%) | 0.02 | 8,062 (51.9%) | <0.01 |
| Alcohol Abuse | 78,884 (3.4%) | 17,943 (3.4%) | 0.67 | 1,173 (7.5%) | <0.01 |
| Drug abuse | 74,146 (3.2%) | 18,161 (3.5%) | 0.03 | 789 (5.1%) | <0.01 |
| **Index presentation** |  |  |  |  |  |
| Heart Failure Decompensations<br>Cardiogenic Shock | 30,328 (1.3%) | 25,117 (4.8%) | <0.01 | 5,165 (33.2%) | <0.01 |
| Cardiac Arrest | 23,310 (1.0%) | 7,572 (1.4%) | <0.01 | 1,262 (8.1%) | <0.01 |
| Mechanical Ventilation | 42,108 (1.8%) | 16,141 (3.1%) | <0.01 | 2,755 (17.7%) | <0.01 |
| Noninvasive Ventilation >24h | 52,098 (2.2%) | 10,946 (2.1%) | 0.29 | 570 (3.7%) | <0.01 |
| Ventricular Arrhythmia | 108,015 (4.6%) | 41,875 (8.0%) | <0.01 | 3,993 (25.7%) | <0.01 |
| Acute Kidney Injury | 667,497 (28.6%) | 183,876 (35.0%) | <0.01 | 9,137 (58.8%) | <0.01 |
| Myocardial Infarction | 109,541 (4.7%) | 29,017 (5.5%) | <0.01 | 3,242 (20.9%) | <0.01 |
| STEMI | 6,671 (0.3%) | 1,910 (0.4%) | <0.01 | 602 (3.9%) | <0.01 |
| Weekend admission | 549,453 (23.6%) | 114,410 (21.8%) | <0.01 | 3,486 (22.4%) | 0.02 |
| Elective admission | 99,739 (4.3%) | 30,274 (5.8%) | <0.01 | 1,051 (6.8%) | <0.01 |
| Primary payer |  |  |  |  |  |
| Medicare | 1,777,430 (76.3%) | 372,486 (71.0%) | <0.01 | 9,212 (59.3%) | <0.01 |
| Medicaid | 210,406 (9.0%) | 56,021 (10.7%) |  | 2,058 (13.2%) |  |
| Private insurance | 237,131 (10.2%) | 71,889 (13.7%) |  | 3,550 (22.8%) |  |
| Self/No Charge/Other | 104,283 (4.5%) | 23,923 (4.6%) |  | 719 (4.6%) |  |
| Disposition status |  |  |  |  |  |
| Home | 1,793,627 (76.9%) | 421,349 (80.3%) | <0.01 | 11,204 (72.1%) | <0.01 |
| Facility | 445,151 (19.1%) | 82,368 (15.7%) |  | 2,493 (16.0%) |  |
| AMA/Unknown/Other | 33,031 (1.4%) | 6,115 (1.2%) |  | 109 (0.7%) |  |
| Died | 59,881 (2.6%) | 15,206 (2.9%) |  | 1,734 (11.2%) |  |

Values are presented as number (percentage) for categorical values and mean (standard error) for continuous variables.
\* Rao-Scott $\chi^2$ test was used for all statistical tests in comparison to Cohort A unless stated otherwise.
† Survey-specific linear regression was performed.

### Characteristics of Patients with All Heart Failure

Patients admitted to ATCs were slightly younger than patients admitted to non-ATCs (69.3 vs 72.5, p < 0.01) with a higher percentage of males (Table 1). The patient comorbidities between these cohorts were generally similar, with patients at ATCs having slightly higher rates of valvular disease, chronic kidney disease, liver disease, malignancy, and coagulopathy, and lower rates of thyroid and pulmonary disease. However, patients treated at ATCs were generally sicker from a HF perspective, with higher rates of HF decompensations: cardiogenic shock (4.8% vs 1.3%, p<0.01), cardiac arrest (1.4% vs 1.0%, p<0.01), mechanical ventilation (3.1% vs 1.8%, p<0.01), ventricular arrhythmias (8.0% vs 4.6%, p<0.01), AKI (35.0% vs 28.6%, p<0.01) (Central Illustration, Table 1).

Patients who were transferred to ATCs (Cohort C) were even sicker, with several-fold higher rates of HF decompensations: cardiogenic shock (33.2% vs 1.3%, p<0.01), cardiac arrest (8.1% vs 1.0%, p<0.01), mechanical ventilation (17.7% vs 1.8%, p<0.01), noninvasive ventilation (3.7% vs 2.2%, p<0.01), ventricular arrhythmias (25.7% vs 4.6%, p<0.01), AKI (58.8% vs 28.6%, p<0.01). These were also much higher compared to Cohort B as well (Supplemental Table 3). A much larger portion of these transfers had myocardial infarction (MI) as the etiology of or coinciding with their HF exacerbation: MI (20.9% vs 4.7%, p<0.01), STEMI (3.9% vs 0.3%, p<0.01). Corresponding with the overall sicker population and increased HF decompensations were also much higher rates of overall comorbidities in patients transferred to ATCs (Table 1). However, transferred patients were younger (63.2 vs 72.5 years, p<0.01) with fewer in the highest age group (14.2% vs 37.6%, p<0.01), less female (39.6% vs 49.2%, p<0.01), and of higher socioeconomic status, with fewer in the lowest geographical income quartile (25.8% vs 33.0%, p<0.01) and higher rates of private insurance (22.8% vs 10.2%, p<0.01).

### Outcomes and Interventions of Patients with All HF

As compared to patients admitted to non-ATCs, patients admitted to ATCs had higher unadjusted mortality (2.9% vs 2.6%, p<0.01), lengths-of-stay, estimated costs, and 90-day readmission rates (Table 2). Patients admitted to ATCs underwent increased rates of HF-related procedures, including right heart catheterization (RHC) (11.8% vs 3.1%, p<0.01) and temporary mechanical circulatory support (MCS) (1.5% vs 0.2%, p<0.01), with the majority being intraaortic balloon pump. Only a minority of these patients received advanced therapies (0.9% LVAD and 0.9% heart transplant).

**Table 2–.** Outcomes/Procedures Performed for Index Heart Failure Admissions, Stratified by Hospital Type and Transfer Status.

|  | **Admission to**<br>**Non-ATC**<br>**(A)** | **Admission to**<br>**ATC**<br>**(B)** | **P value\*** | **Transfer to**<br>**ATC**<br>**(C)** | **P value\*** |
| --- | --- | --- | --- | --- | --- |
|  | n = 2,331,690 | n = 525,037 |  | n = 15,541 |  |
| Mortality | 59,881 (2.6%) | 15,206 (2.9%) | <0.01 | 1,734 (11.2%) | <0.01 |
| Length of Stay (days) | 4.9 (SEM = 0.01) | 6.6 (SEM = 0.07) | <0.01<sup>†</sup> | 14.6 (SEM = 0.25) | <0.01<sup>†</sup> |
| Estimated Cost (\$) | 11,046<br>(SEM = 46.92) | 19,263<br>(SEM = 435.79) | <0.01<sup>†</sup> | 57,092<br>(SEM = 1896.15) | <0.01<sup>†</sup> |
| 90-day readmission in survivors | 835,103 (36.8%) | 194,167 (38.1%) | <0.01 | 5,197 (37.6%) | 0.19 |
| Procedures Performed |  |  |  |  |  |
| RHC | 73,406 (3.1%) | 61,713 (11.8%) | <0.01 | 6,950 (44.7%) | <0.01 |
| PCI | 26,030 (1.1%) | 7,820 (1.5%) | <0.01 | 1,105 (7.1%) | <0.01 |
| CABG | 3,863 (0.2%) | 1,975 (0.4%) | <0.01 | 338 (2.2%) | <0.01 |
| Temporary mechanical support | 4,384 (0.2%) | 7,934 (1.5%) | <0.01 | 2,190 (14.1%) | <0.01 |
| IABP | 2,426 (0.1%) | 4,145 (0.8%) | <0.01 | 1,269 (8.2%) | <0.01 |
| PVAD | 1,991 (<0.1%) | 1,780 (0.3%) | <0.01 | 757 (4.9%) | <0.01 |
| ECMO | 87 (<0.1%) | 1,467 (0.3%) | <0.01 | 579 (3.7%) | <0.01 |
| TAVR | 706 (<0.1%) | 752 (0.1%) | <0.01 | 68 (0.4%) | <0.01 |
| LVAD implantation | - | 4,485 (0.9%) | - | 549 (3.5%) | - |
| Heart Transplantation | - | 4,785 (0.9%) | - | 116 (0.7%) | - |

Values are presented as number (percentage) for categorical values and mean (standard error) for continuous variables. CABG indicates coronary artery bypass graft; ECMO, extracorporeal membrane oxygenation; IABP, intraaortic balloon pump; LVAD, left ventricular assist device; PCI, percutaneous coronary intervention; PVAD, percutaneous ventricular assist device; RHC, right heart catheterization; TAVR, transcatheter aortic valve replacement.
\* Rao-Scott $\chi^2$ test was used for statistical tests in comparison to Cohort A unless stated otherwise.
<sup>†</sup> Survey-specific linear regression was performed.

Patients transferred to ATCs had even higher unadjusted mortality (11.2% vs 2.6%, p<0.01), lengths-of-stay, and estimated costs, while readmission rates were similar. Transferred patients underwent a dramatically higher rate of HF-related procedures, including RHC (44.7% vs 3.1%, p<0.01), percutaneous coronary intervention (PCI) (7.1% vs 1.1%, p<0.01), and MCS (14.1% vs 0.2%, p<0.01). Compared to patients directly admitted to ATCs, patients transferred to ATCs underwent a higher rate of LVAD implantation (3.5% vs 0.9%, p<0.01), and a similar rate of heart transplantation (0.7% vs 0.9%, p = 0.27) (Supplemental Table 4).

Multivariable analysis was performed to account for differences in comorbidities and rates of HF decompensations among the groups. Despite the higher unadjusted mortality, the adjusted mortality was significantly lower at ATCs, both for directly admitted (OR 0.82, p<0.01) and transferred patients (OR 0.66, p<0.01) (Table 3). Each HF decompensation studied other than ventricular arrhythmias conferred a several-fold higher odds of mortality (Table 3).

**Table 3–.** Multivariable Analysis for Predictors of Mortality – Index Heart Failure Admissions.

| **Characteristic** | **Odds Ratio**<br>**(95% Confidence**<br>**Interval)** | **P value** |
| --- | --- | --- |
| B vs A | 0.82 (0.78-0.87) | <0.01 |
| C vs A | 0.66 (0.57-0.78) | <0.01 |
| C vs B | 0.81 (0.70-0.94) | <0.01 |
| **Index Admission Characteristics** |  |  |
| Elective Admission | 1.19 (1.05-1.36) | <0.01 |
| Weekend Admission | 0.97 (0.94-1.00) | 0.05 |
| Myocardial Infarction | 1.72 (1.64-1.81) | <0.01 |
| **Heart Failure Decompensations** |  |  |
| Cardiogenic Shock | 6.20 (5.86-6.57) | <0.01 |
| Cardiac Arrest | 22.25 (20.70-23.91) | <0.01 |
| Mechanical Ventilation | 9.19 (8.74-9.66) | <0.01 |
| Noninvasive Ventilation >24h | 3.49 (3.30-3.69) | <0.01 |
| Ventricular Arrhythmia | 0.86 (0.81-0.91) | <0.01 |
| Acute Kidney Injury | 2.19 (2.12-2.26) | <0.01 |
| **Patient Comorbidities** |  |  |
| Age* | 1.67 (1.64-1.70) | <0.01 |
| Female sex | 0.99 (0.96-1.01) | 0.30 |
| Hypertension | 0.88 (0.85-0.91) | <0.01 |
| Hyperlipidemia | 0.71 (0.68-0.73) | <0.01 |
| Coronary Artery Disease | 0.93 (0.90-0.96) | <0.01 |
| Diabetes | 0.90 (0.87-0.92) | <0.01 |
| Atrial Fibrillation/Flutter | 1.28 (1.24-1.32) | <0.01 |
| Valvular Disease | 0.95 (0.92-0.98) | <0.01 |
| Peripheral vascular disease | 1.08 (1.04-1.13) | <0.01 |
| Chronic Lung Disease | 1.07 (1.04-1.10) | <0.01 |
| Chronic Kidney Disease | 1.05 (1.02-1.08) | <0.01 |
| Liver Disease | 1.43 (1.36-1.50) | <0.01 |
| Malignancy | 1.54 (1.47-1.61) | <0.01 |
| Smoking History | 0.79 (0.77-0.82) | <0.01 |
| Alcohol Abuse | 0.78 (0.71-0.86) | <0.01 |
| Drug abuse | 0.73 (0.65-0.82) | <0.01 |
| Thyroid Disease | 1.03 (1.00-1.07) | 0.05 |
| Autoimmune Disease | 1.11 (1.04-1.19) | <0.01 |
| Coagulopathy | 1.38 (1.33-1.44) | <0.01 |
| Chronic Anticoagulation | 0.76 (0.73-0.79) | <0.01 |
| Obesity | 0.78 (0.75-0.81) | <0.01 |
| Malnutrition | 2.08 (2.00-2.17) | <0.01 |
| Median household income by zip code<br>(quartile) |  |  |
| 1<sup>st</sup> | 1 (reference) | - |
| 2<sup>nd</sup> | 1.13 (1.09-1.18) | <0.01 |
| 3<sup>rd</sup> | 1.10 (1.06-1.15) | <0.01 |
| 4<sup>th</sup> | 1.09 (1.04-1.14) | <0.01 |
| Primary Payer<br>Medicare | 1 (reference) | - |
| Medicaid | 0.81 (0.76-0.87) | <0.01 |
| Private insurance | 0.92 (0.87-0.98) | <0.01 |
| Self/No Charge/Other | 1.13 (1.04-1.23) | <0.01 |

\* Per increase in age group by 10 years

**Table 4–.** Prevalence and Mortality Associated with Heart Failure Decompensations During All Index Admissions for Heart Failure.

| **Heart Failure Decompensations** | **Prevalence** | **Mortality** |
| --- | --- | --- |
| Cardiogenic Shock | 60,610 (2.1%) | 27.9% |
| Cardiac Arrest | 32,144 (1.1%) | 58.3% |
| Mechanical Ventilation | 61,004 (2.1%) | 40.2% |
| Noninvasive Ventilation >24h | 63,614 (2.2%) | 8.6% |
| Ventricular Arrhythmias | 153,883 (5.4%) | 7.5% |
| Acute Kidney Injury | 860,511 (30.0%) | 5.3% |

Given that non-ATCs represent a large heterogenous group of hospitals, a sensitivity analysis was performed to include only CABG-capable hospitals, which showed similar findings (Supplemental Table 9).

### Characteristics of Patients with Severe HF

A secondary analysis was performed for the subset of patients hospitalized with severe HF to isolate this group from the broader population with milder HF, given the higher mortality rates associated with HF decompensations: cardiogenic shock, cardiac arrest, or mechanical ventilation (Supplemental Table 5). There were 89,843 (61.5%) patients with severe HF admitted to non-ATCs, 48,488 (33.2%) admitted to ATCs, and 7,825 (5.4%) transferred to ATCs (Table 5).

**Table 5-.**
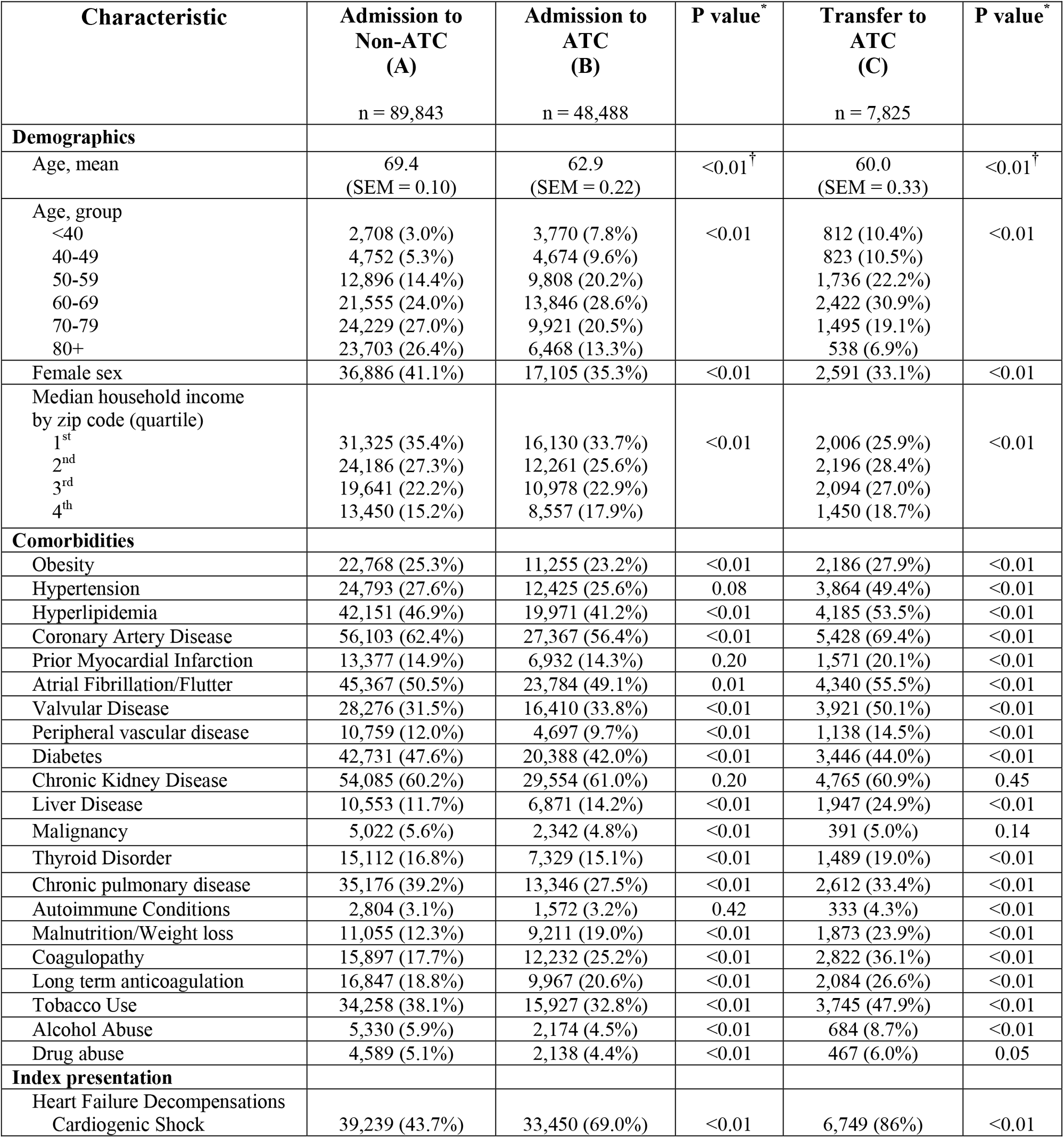
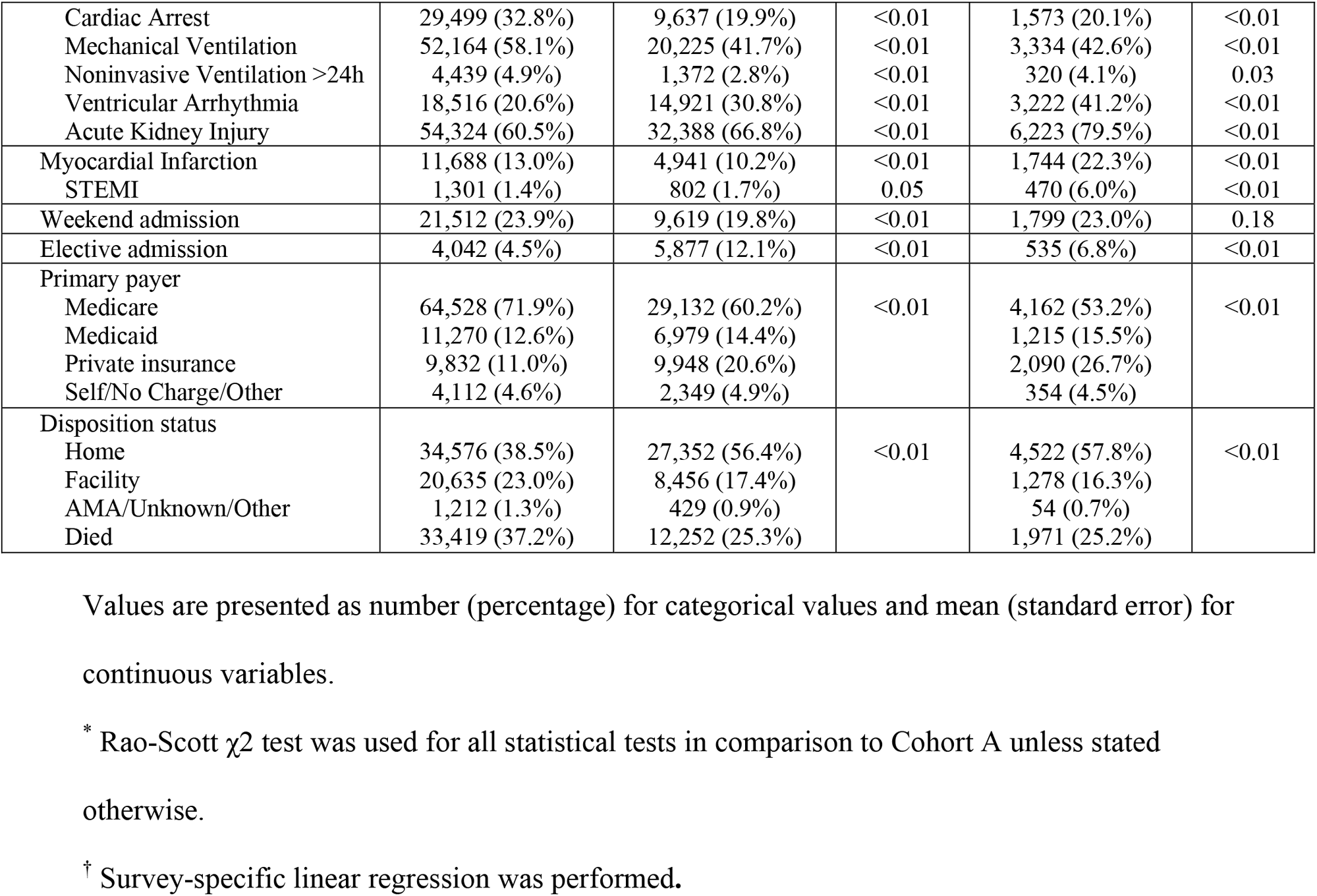
Characteristics of Patients Admitted with Severe Heart Failure, Stratified by Hospital Type and Transfer Status.

Compared to patients admitted to non-ATCs, patients at ATCs with severe HF were significantly younger (62.9 vs 69.4 years, p<0.01) and male-predominant (Table 5). Patients at ATCs had higher rates of some comorbidities, including valvular disease, liver disease, malnutrition/weight loss, coagulopathy, and lower rates of other comorbidities, including hyperlipidemia, coronary artery disease, peripheral arterial disease, diabetes, malignancy, and chronic lung disease. Patients who were transferred, similar to the all-comer HF population, had higher rates of most comorbidities. Similar to the all-comer HF population, transferred patients were notably younger (Age 60.0 vs 69.4, p<0.01) with less females (33.1% vs 41.1%, p<0.01), and reflected a higher sociodemographic status, with significantly fewer in the lowest geographical income quartile (25.9% vs 35.4%, p<0.01), and higher rates of private insurance (26.7% vs 11.0%, p<0.01).

### Outcomes and Interventions of Patients with Severe HF

For patients with severe HF, in-hospital mortality was notably lower at ATCs, whether patients were directly admitted (25.3% vs 37.2%, p<0.01) or transferred (25.2% vs 37.2%, p<0.01), although corresponding to higher costs and lengths-of-stay, especially for transferred patients (Table 6). HF-related procedures were performed at a much higher rate in general for this severe HF population, with higher procedure rates at ATCs, including RHC (41.4% vs 9.8%, p<0.01) and MCS (17.4% vs 3.7%, p<0.01). Higher numerical rates of LVAD (10.1%) and transplant (7.2%) also occurred for this sicker population as compared to the general HF population. For severe HF patients who were transferred to ATCs, the HF-related procedures were performed at an even higher rate, with almost two thirds of patients receiving RHC (61.8% vs 9.8%, p<0.01) and almost one third receiving MCS (31.7% vs 3.7%, p<0.01). However, the incidence of LVAD or transplant was not higher for patients transferred than for patients directly admitted to ATCs (Table 6, Supplemental Table 6).

**Table 6–.**
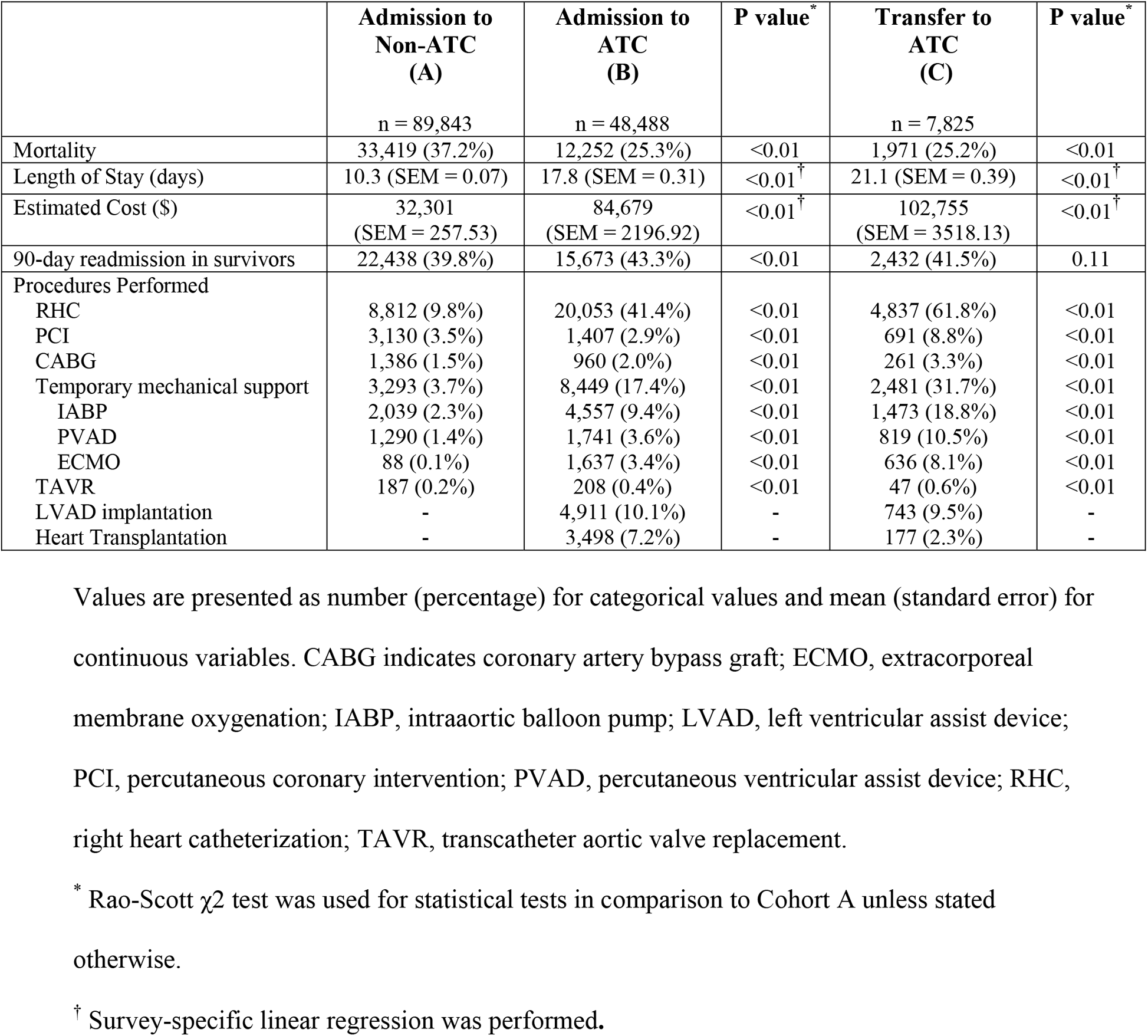
Outcomes/Procedures Performed for Index Severe Heart Failure Admissions, Stratified by Hospital Type and Transfer Status.

|  | **Admission to**<br>**Non-ATC**<br>**(A)**<br>n = 89,843 | **Admission to**<br>**ATC**<br>**(B)**<br>n = 48,488 | **P value\*** | **Transfer to**<br>**ATC**<br>**(C)**<br>n = 7,825 | **P value\*** |
| --- | --- | --- | --- | --- | --- |
| Mortality | 33,419 (37.2%) | 12,252 (25.3%) | <0.01 | 1,971 (25.2%) | <0.01 |
| Length of Stay (days) | 10.3 (SEM = 0.07) | 17.8 (SEM = 0.31) | <0.01<sup>†</sup> | 21.1 (SEM = 0.39) | <0.01<sup>†</sup> |
| Estimated Cost (\$) | 32,301<br>(SEM = 257.53) | 84,679<br>(SEM = 2196.92) | <0.01<sup>†</sup> | 102,755<br>(SEM = 3518.13) | <0.01<sup>†</sup> |
| 90-day readmission in survivors | 22,438 (39.8%) | 15,673 (43.3%) | <0.01 | 2,432 (41.5%) | 0.11 |
| Procedures Performed |  |  |  |  |  |
| RHC | 8,812 (9.8%) | 20,053 (41.4%) | <0.01 | 4,837 (61.8%) | <0.01 |
| PCI | 3,130 (3.5%) | 1,407 (2.9%) | <0.01 | 691 (8.8%) | <0.01 |
| CABG | 1,386 (1.5%) | 960 (2.0%) | <0.01 | 261 (3.3%) | <0.01 |
| Temporary mechanical support | 3,293 (3.7%) | 8,449 (17.4%) | <0.01 | 2,481 (31.7%) | <0.01 |
| IABP | 2,039 (2.3%) | 4,557 (9.4%) | <0.01 | 1,473 (18.8%) | <0.01 |
| PVAD | 1,290 (1.4%) | 1,741 (3.6%) | <0.01 | 819 (10.5%) | <0.01 |
| ECMO | 88 (0.1%) | 1,637 (3.4%) | <0.01 | 636 (8.1%) | <0.01 |
| TAVR | 187 (0.2%) | 208 (0.4%) | <0.01 | 47 (0.6%) | <0.01 |
| LVAD implantation | - | 4,911 (10.1%) | - | 743 (9.5%) | - |
| Heart Transplantation | - | 3,498 (7.2%) | - | 177 (2.3%) | - |

Values are presented as number (percentage) for categorical values and mean (standard error) for continuous variables. CABG indicates coronary artery bypass graft; ECMO, extracorporeal membrane oxygenation; IABP, intraaortic balloon pump; LVAD, left ventricular assist device; PCI, percutaneous coronary intervention; PVAD, percutaneous ventricular assist device; RHC, right heart catheterization; TAVR, transcatheter aortic valve replacement.
\* Rao-Scott $\chi^2$ test was used for statistical tests in comparison to Cohort A unless stated otherwise.
<sup>†</sup> Survey-specific linear regression was performed.

Multivariable analysis also resulted in lower adjusted mortality for patients with severe HF admitted (OR 0.81, p<0.01) or transferred to ATCs (OR 0.77, p<0.01) (Table 7) that corresponded to the lower unadjusted mortality. The mortality differences for both HF groups were present even after further adjustment for procedural use (Supplemental Table 7 and 8).

**Table 7–.** Multivariable Analysis for Predictors of Mortality – Index Severe Heart Failure Admissions.

| **Characteristic** | **Odds Ratio**<br>**(95% Confidence**<br>**Interval)** | **P value** |
| --- | --- | --- |
| B vs A | 0.81 (0.76-0.85) | <0.01 |
| C vs A | 0.77 (0.69-0.86) | <0.01 |
| C vs B | 0.96 (0.86-1.06) | 0.40 |
| **Index Admission Characteristics** |  |  |
| Elective Admission | 0.75 (0.69-0.81) | <0.01 |
| Weekend Admission | 0.97 (0.93-1.02) | 0.20 |
| Myocardial Infarction | 1.15 (1.09-1.22) | <0.01 |
| **Heart Failure Decompensations** |  |  |
| Cardiogenic Shock | 1.93 (1.83-2.03) | <0.01 |
| Cardiac Arrest | 7.66 (7.25-8.08) | <0.01 |
| Mechanical Ventilation | 2.96 (2.83-3.11) | <0.01 |
| Noninvasive Ventilation >24h | 1.52 (1.38-1.67) | <0.01 |
| Ventricular Arrhythmia | 0.64 (0.60-0.67) | <0.01 |
| Acute Kidney Injury | 1.31 (1.25-1.37) | <0.01 |
| **Patient Comorbidities** |  |  |
| Age | 1.36 (1.33-1.39) | <0.01 |
| Female sex | 0.98 (0.94-1.02) | 0.35 |
| Hypertension | 0.89 (0.85-0.93) | <0.01 |
| Hyperlipidemia | 0.82 (0.79-0.85) | <0.01 |
| Coronary Artery Disease | 0.95 (0.91-0.99) | <0.01 |
| Diabetes | 0.96 (0.92-1.00) | 0.03 |
| Atrial Fibrillation/Flutter | 1.05 (1.00-1.09) | 0.03 |
| Valvular Disease | 0.96 (0.92-1.00) | 0.06 |
| Peripheral vascular disease | 1.13 (1.06-1.19) | <0.01 |
| Chronic Lung Disease | 0.94 (0.90-0.97) | <0.01 |
| Chronic Kidney Disease | 1.07 (1.03-1.12) | <0.01 |
| Liver Disease | 1.26 (1.19-1.34) | <0.01 |
| Malignancy | 1.40 (1.29-1.51) | <0.01 |
| Smoking History | 0.88 (0.85-0.92) | <0.01 |
| Alcohol Abuse | 0.71 (0.64-0.78) | <0.01 |
| Drug abuse | 0.70 (0.64-0.78) | <0.01 |
| Thyroid Disease | 1.04 (0.99-1.10) | 0.15 |
| Autoimmune Disease | 1.13 (1.02-1.26) | 0.02 |
| Coagulopathy | 1.31 (1.25-1.37) | <0.01 |
| Chronic Anticoagulation | 0.93 (0.88-0.98) | <0.01 |
| Obesity | 0.85 (0.82-0.89) | <0.01 |
| Malnutrition | 1.14 (1.08-1.20) | <0.01 |
| Median household income by zip code<br>(quartile) |  |  |
| 1<sup>st</sup> | 1 (reference) | - |
| 2<sup>nd</sup> | 1.06 (1.01-1.12) | 0.02 |
| 3<sup>rd</sup> | 1.08 (1.03-1.14) | <0.01 |
| 4<sup>th</sup> | 1.05 (0.99-1.12) | 0.09 |
| Primary Payer |  |  |
| Medicare | 1 (reference) | - |
| Medicaid | 0.79 (0.74-0.85) | <0.01 |
| Private insurance | 0.81 (0.76-0.86) | <0.01 |
| Self/No Charge/Other | 0.99 (0.89-1.09) | 0.77 |

\* Per increase in age group by 10 years

## Discussion

To our knowledge, this is the largest contemporary study to examine the characteristics of HF admissions and transfers as they relate to ATCs. Our main findings are: (1) patients admitted to ATCs were generally sicker with more HF decompensations and higher procedural volume, lengths-of-stay, cost, and unadjusted mortality, (2) when accounting for disease severity and decompensations, adjusted mortality for HF admissions was lower at ATCs, (3) patients transferred to ATCs were even sicker and associated with several-fold higher rates of HF decompensations, procedural volume, and correspondingly higher unadjusted mortality; however adjusted mortality was also lower.

Much of the focus of study in recent years has been on cardiogenic shock as an intensive, rapidly progressive, highly-morbid disease state necessitating a multidisciplinary approach, regional networks, and hub-and-spoke models to enable the early and rapid referral or transfer of critically-ill patients to centralized hubs in order to improve outcomes^8–11^. However, shock is the last stage in the spectrum of acute decompensated HF, and earlier phases of this disease process are also associated with significant morbidity and mortality^3,5^. The gold standard of HF care remains at centers that can provide the highest level of advanced HF therapies, such as LVAD or transplant, as well as other non-advanced HF therapies and resources. Although there are a variety of tools used to guide appropriate referrals to higher level care, ranging from functional class, biomarkers, clinical markers, and hemodynamics, appropriate and timely referral to an ATC likely continues to be lacking^5,12^. Furthermore, there is also a paucity of contemporary data that examines the patterns of HF treatment at different center types and the role and impact of ATCs on outcomes of HF treatment.

Our study showed that ATCs formed a small percentage of all hospitals and treated a minority (but still a disproportionally higher percentage) of HF admissions. When admitted for HF, patients at advanced therapy centers were sicker with a significantly higher burden of decompensations. While this may reflect a concentration of more advanced HF patients who were already under the umbrella of these ATCs, this may also reflect under-diagnosis of more severe HF at non-ATCs. For example, patients with progressive HF may not be recognized as an outpatient until they are admitted in the throes of a decompensation, and even when admitted, comorbidities, low output, impending respiratory compromise, and other decompensations may be underrecognized or recognized late. Notably, when adjusting for disease severity either with multivariable analysis or by isolating the severe HF cohort, HF admissions at ATCs were associated with lower mortality within our study.

There are likely multiple reasons for the lower mortality at ATCs seen in our study. Patients at ATCs were treated with significantly increased numbers of HF-related procedures, beyond just LVAD and transplant. RHC in particular was frequently performed, which has been shown to be beneficial in the evaluation and management of cardiogenic shock^11,15,16^. Various forms of temporary MCS were also used more frequently at advanced therapy centers, and particularly for transferred patients. Temporary MCS may be used for a variety of reasons in HF, including as a bridge to recovery, intervention, or advanced therapies. Studies on the use of temporary MCS in the sicker cardiogenic shock population have been limited and conflicting, perhaps reflecting the complexity of MCS, the overall severity of illness in patients who require MCS, and the need to be tailored to specific clinical situations and hemodynamics^7,8,16,17^. In the case of transfer specifically, potential uses of temporary MCS may include stabilization and support of patients to allow for transfer, or as part of target interventions at the ATC. Finally, other therapeutic procedures such as PCI, CABG, and TAVR were also utilized more frequently at ATCs and may also represent target interventions in transferred patients. The higher rates of heart-failure related procedural use and availability may play a role in the improved outcomes seen at ATCs within our study.

Although ATCs were defined based on capability of advanced therapies, the lower associated mortality is observed even though the percentage of patients actually receiving an LVAD or transplant were each less than 1%. Furthermore, lower associated mortality remained even after additional adjustment for procedural use, suggesting that the non-procedural resources at ATCs can also contribute to improved outcomes. Such resources may include dedicated cardiac floors and cardiac care units, equipment, improved nursing ratios, educational resources, staff more experienced in recognizing and treating HF decompensations, and the availability of more experienced multidisciplinary practitioners such as dedicated HF physicians, interventional cardiologists, electrophysiologists, cardiac surgeons, and critical care specialists. Of note, non-ATCs still represent a wide range of centers and capabilities, and future studies dedicated towards the impact of different levels of non-ATCs will be useful.

Notably, patients who were transferred to ATCs represented by far the sickest population, with the highest rates of HF decompensations and comorbidities. However, despite the disease severity, the observed adjusted mortality was lower. These patients had several-fold higher rates of procedural frequency than both non-transferred cohorts. In fact, many may be transferred specifically with therapeutic interventions in mind (revascularization, MCS, LVAD, etc.).

Interestingly, transferred patients were younger, male-predominant, and with higher socioeconomic status. On one hand, this likely represents a selection bias that parallels real-world selection processes in clinical practice, possibly reflecting a desire to transfer younger and more “viable” patients (despite being sicker) with socioeconomic support and promising candidacy for interventions including advanced therapies, while screening out patients who are older or too sick, for the best overall use of limited resources. Multidisciplinary patient selection processes play important roles in advanced therapy selection meetings in order to best allocate scarce resources, and may play a similar role in the transfer screening process for HF patients. On the other hand, this may represent a disparity to care for certain patient populations with HF, where older age groups, women, and patients with lower socioeconomic status do not have access to the same level of attention or resources. A variety of factors could contribute to this disparity at the patient, provider, financial, and/or geographical level. Both socioeconomic and gender disparities have been previously demonstrated in the field of cardiology^18–22^. Further study of the decision-making and selection process for transfers, and its impact with regards to healthcare disparities, is needed.

Finally, although mortality was lower at ATCs both for directly admitted and transferred patients, not all HF admissions can realistically be treated at ATCs. There is a role for increased ATC utilization, as even for the severe HF admissions, 37.2% of patients were dying at non-ATCs with low percentages of possibly beneficial HF-related procedures, rather than being transferred to ATCs. However, it is notable that ATCs represented only 5.5% of all hospitals in our study, yet treated nearly 20% of all HF patients. In an ideal world, all patients would have access to the highest resource concentration and expertise at ATCs, but given the significantly higher costs and lengths-of-stay associated with the care or transfer to ATCs and the low number of ATCs available, this ideal is unlikely to be feasible in the near future. Possible solutions may involve exploring further avenues to extend the multidisciplinary expertise at ATCs, including virtual consults, cross-institutional relationship building, and ATC provider outreach/presence at non-ATC centers in order to share expertise and experience, encourage prevention, and facilitate transfers. Additionally, while efforts to improve resources and recognition at non-ATCs are important, the aforementioned screening/selection process as part of a multidisciplinary team will continue to be important to maximize the benefit/cost balance in order to provide the best HF care for the most patients possible. Further studies evaluating the cost/benefit relationship as it pertains to patient referral to ATCs are needed.

The strength of our study was the use of a large, inclusive, real-world sample of HF hospitalizations to assess patterns of HF admissions and transfers and the impact of ATCs on a national level. We also acknowledge several limitations. First, our data is observational, retrospective, and derived from a national administrative database, and inaccurate coding is a fundamental limitation of any database based on diagnosis codes. HCUP routinely performs quality control procedures to verify validity of NRD data^14,23^. However, there could also be differences in patterns of coding among types of institutions (i.e. ATCs may code more comprehensively to capture their patient complexity). Second, the NRD is a national database combined from state inpatient databases, and a small proportion of transfers nationally will not be recognized in the NRD (transfer across different states, or involving federal hospitals). Third, the origin of the transfer is not included, so a small proportion may represent inter-ATC transfer. Fourth, the NRD does not contain more detailed patient data including lab values, organ function, hemodynamics, and reason and timing of transfer, which limits insight into the decision-making and selection process during admission or transfer. Finally, as a national inpatient database, the NRD does not contain data on outpatient patterns of referral and longer-term outcomes after hospitalization. Further study will be needed to assess the impact of these factors on admission and transfer patterns, outcomes, and role of ATCs in HF admissions.

## Data Availability

The data that support the findings of this study are available from the corresponding author upon reasonable request.

https://hcup-us.ahrq.gov/nrdoverview.jsp?_gl=1*ffpf55*_ga*MTI4OTg5OTQ0LjE2ODg3NzUwMDU.*_ga_1NPT56LE7J*MTY5MTI3MTMzMC4zLjEuMTY5MTI3MTMzNi41NC4wLjA.

## Non-standard Abbreviations and Acronyms

ATC: Advanced Therapy Center
NRD: National Readmissions Database
AHRQ: Agency for Healthcare Research and Quality
HCUP: Healthcare Cost and Utilization Project
SID: State Inpatient Databases

## Acknowledgements

None

## Sources of Funding

This work was supported by grants from the Michael Wolk Heart Foundation (New York, NY), the New York Cardiac Center, Inc (New York, NY), and the New York Weill Cornell Medical Center Alumni Council (New York, NY). The Michael Wolk Heart Foundation, the New York Cardiac Center, Inc, and the New York Weill Cornell Medical Center Alumni Council had no role in the design and conduct of the study, in the collection, analysis, and interpretation of the data, or in the preparation, review, or approval of the manuscript.

## Disclosures

Dr. Cheung has received consulting fees from Abbott, Boston Scientific and Biotronik, fellowship grant support from Abbott, Biotronik, Boston Scientific, and Medtronic, and research support from Boston Scientific. Dr. Minutello is on the advisory board for Medtronic. Dr. Goyal is supported by the American Heart Association grant 18IPA34170185 and is a recipient of a National Institute on Aging Loan Repayment Plan. Dr. Horn has received consulting income from Biotronik. Dr. Uriel is on the advisory board of Livemetric, Revamp, and Leviticus, and has received grant support from Abbott and Abiomed. Dr. Kim has received consulting fees from Axon Therapies and fellowship grant support from Medtronic and Abbott. All other authors have reported no conflict of interest relevant to the publication of this paper.

## Supplemental Material

Tables S1 – S9

## Notes

### Competing Interest Statement

The authors have declared no competing interest.

### Author Declarations

Given all data were derived from a de-identified national administrative database, Institutional Review Board (IRB) approval and informed consent were not required.

